# Geospatial approach to investigate spatial clustering and hotspots of blood lead levels in children within Kabwe, Zambia

**DOI:** 10.1101/2021.03.16.21253682

**Authors:** Given Moonga, Moses N Chisola, Ursula Berger, Dennis Nowak, John Yabe, Hokuto Nakata, Shouta Nakayama, Mayumi Ishizuka, Stephan Bose-O’Reilly

## Abstract

**Background:** Communities around Kabwe/Zambia are exposed to lead due to deposits from an old lead (Pb) and zinc (Zn) mining site. Children are particularly more vulnerable than adults, presenting with greatest risk of health complications because of their increased oral uptake due to their hand to mouth activities. Spatial analysis of childhood lead exposure is useful in identifying specific areas with highest risk of pollution. The objective of the current study was to use a geospatial approach investigate spatial clustering and hotspots of blood lead levels in children within Kabwe.

**Methods:** We analysed data on blood lead levels (BLL) for 363 children below the age of 15 from Kabwe town. We used spatial autocorrelation methods involving the global Moran’s I and local Getis-Ord Gi*statistic in ArcMap 10.5.1, to test for spatial dependency among the blood lead levels in children.

**Results:** BLL in children from Kabwe are spatially autocorrelated with a Moran’s Index of 0.62 (p<0.001). We found distinct hot spots in communities close to the old lead and zinc-mining site, lying on its western side. We observed lower levels of BLL in areas distant to the mine and located at its eastern side. This pattern suggests a possible association between BLL and distance from the abandoned lead and zinc mine, and prevailing winds.

**Conclusion:** Using geocoded data, we found clustering of childhood blood lead and identified distinct hot spot areas with particular high lead levels for Kabwe town. The geospatial approach used is especially valuable in resource-constrained settings like Zambia, where the precise identification of the locations of risk areas allows to initiate targeted remedial and treatment programs.

## 1. Introduction

Lead (Pb) is a toxic metal and a global health hazard. Over 815 million children worldwide are reported to have dangerously high concentrations of Pb in their bloodstream (Burki, 2020) with low-income countries of sub-Saharan Africa facing the greatest impacts of childhood lead poisoning (Landrigan et al., 2018). Children are more vulnerable to Pb related negative health outcomes, compared to adults, as their still developing central nervous system is more subtle to Pb exposure, mainly during the primary developmental stages (Rooney et al., 2018, Scheuplein et al., 2002). Pb exposure, even at low levels, has been associated with deficits in cognitive functioning and intelligence quotient (IQ) in children (Liu et al., 2011, Lanphear et al., 2005). No level of Pb exposure appears to be safe (Bellinger et al., 1991). High Pb levels of exposure exceeding 80 μg/dL can lead to anaemia, seizures, coma, encephalopathy, and death (World Health Organization, 2010, Wang et al., 2009)

While the physiological mechanisms for Pb dose–response may be similar for all children (Mielke et al., 2019), there are varied exposure pathways. The blood lead levels (BLL) are related to Pb soil contamination (Matte et al., 1991) and social economic factors (Mielke et al., 2019). The impact of socio-economic and environmental covariates on the Pb exposure in children has been well documented (Stark et al., 1982, Mielke et al., 2019).

Globally childhood Pb poisoning remains a major environmental health concern in cities and communities with Pb-contaminated soils (Ikem et al., 2008, Lo et al., 2012). Contaminated soil, especially if the soil is dry and dusty, can be ingested and absorbed. Children are particularly vulnerable to Pb intoxication due to their very relevant hand-to-mouth activities they ingest more Pb from contaminated soils, and also due to their much higher gastrointestinal absorption relatively to adults (World Health Organization, 2010, Plumlee et al., 2013).

Kabwe town, located in the Central Province of Zambia had a long history of lead (Pb), zinc (Zn) and cadmium (Cd) mining dating back to the 1900’s. The operations came to a stop in 1994, leaving behind ineliminable impact of pollution on the environment (Nakayama et al., 2011). The open cast mining and the resulting big tailing hill both still contain a lot of Pb. Neither the tailing hill nor the open pit side were ever properly rehabilitated. Since decades, especially during the 9 month period of dry season, Pb can be easily transported into the windward lying areas and communities. Most roads are unpaved, and esp. during the dry season the backyards of the housing areas and dry and dusty.

Ettler et al. (2020) analysed soil samples from Kabwe townships and main roads and reported that soil Pb levels were above recommended levels for residential areas (ATSDR, 2016). They also observed that the geometric mean soil Pb in townships closer to the mining sites were higher than far off areas. Highly polluted townships were those immediately adjacent to the former Kabwe mining complex and homes downwind from the smelter and the tailings (Bose-O’Reilly, Yabe et al. 2018).

Former reports and scientific publications showed increased and even high BLL for people living in Kabwe, due to their continued exposure to lead. Many children in Kabwe residential areas are reported to have BLL of above 65 μg/dL(Yabe et al., 2015a, Yabe et al., 2020). The Pb contaminated dust, that emanates from the mine dump, seems to be the main source, according to the former results. Children are the most affected (Bose-O’Reilly et al., 2018, Yabe et al., 2020, Yabe et al., 2015a).

### Spatial patterns of Pb exposure in Kabwe

Understanding spatial patterns of the Pb burden is particularly important to identify areas with high risk of exposure. (Akkus and Ozdenerol, 2014, Miranda et al., 2002). In resource-limited settings such as Zambia, use of Geographic Information System (GIS) tools to identify affected populations is essential in ensuring that remediation and treatment programmes target those at greatest risk. The integration of GIS to understand Pb exposure patterns and coverage of interventions also strengthens the implementation of control programmes. Spatial analytical methods such as cluster and hot spot analysis are ideal in identifying spatial patterns of exposure or disease in communities (Zhang et al., 2008, Akkus and Ozdenerol, 2014). Moreover, hotspot analysis can help understand disparities in exposure and health outcomes at a lower administrative level. This precision is valuable as it enables quantification of inequalities and identification of successes and failures of programmes and policies at the local level (Osgood-Zimmerman et al., 2018).

Previous studies observed very high BLL among children in Kabwe that varied depending on the townships (Yabe et al., 2020, Bose-O’Reilly et al., 2018). Recent work also observed these regional differences in BLL, and further established that BLL was dependent on distance from the mine site. It was apparent that residential areas (Kasanda, Makululu, Chowa) closer to the mine site had higher corresponding BLL while further located areas (Hamududu) had lower BLL (Yabe et al., 2020). Another study also observed this trend in domestic dogs. The blood Pb concentration was higher in dogs from communities that were located near the mine than the far-flung residential areas (Toyomaki et al., 2020).

Despite the indication of regional variation in the distribution of BLL in Kabwe, until now no study has investigated the spatial distribution of BLL using geospatial techniques to identify hot spot areas. Hot spots in this context are areas with high Pb values surrounded by observation with high values (spatial clusters). As such, observation with a high value or low value does not necessarily imply a hot spot or a cold spot respectively, unless it is surrounded by observations with high values (hot spot) or low values (cold spot). The aim of this study therefore, was to apply GIS tools on BLL in children below the age of 15 in order to investigate clustering and identify hot spot communities within Kabwe.

## 2. Methodology

### 2.1 Study population

We analysed BLL data that was collected in 2017 by the University of Zambia with collaborators from Hokkaido University under the Kabwe Mine Pollution Amelioration Initiative (KAMPAI) Project. Forty (40) Standard Enumeration Areas (SEA) falling within the catchment area of health facilities were randomly selected from which 25 households each were randomly selected. From each household, blood was collected from the father, mother and two children including the geo-coordinates for each household. Data was collected on yx households with a total of 1190 household members, of which 291 were younger children (three months to three years old), 271 older children (four to nine years old), 412 mothers, and 216 fathers. A detailed description on how the data was collected has been provided by Yabe et al. (2020).

We analysed data for 363 children below the age of 15 from the 40 SEAs. We selected the youngest child from each household that was enlisted in the study by Yabe et al. (2020). We focus our analysis on children as they are reported to be the most vulnerable to impacts of lead poisoning. (Bose-O’Reilly et al., 2018). Moreover, this is the age range reported to have highest BLL. Figure 1, shows the residential areas from which households were included in current study.

**Figure 1:**
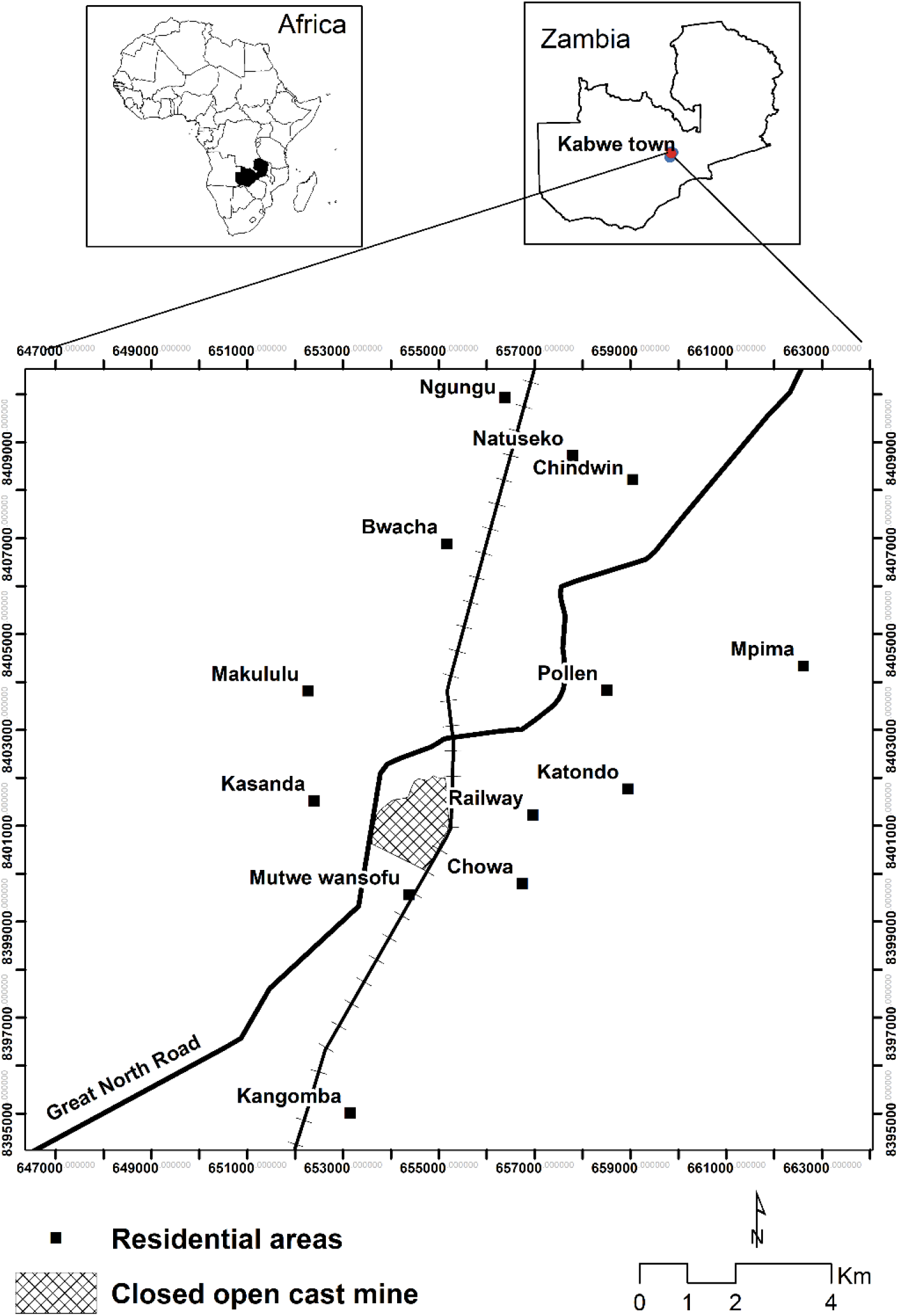
Study area

### 2,2. Laboratory methods

Pb analysis in whole blood samples was done on-site immediately after blood sample collection using a point-of-care blood Pb testing analyser, LeadCare© II (Magellan Diagnostics, USA). The LeadCare II Analyser used had limit of quantification of 3.3 to 65 µg/dL, as such, precise levels out of this range could not be determined. Detailed laboratory procedures are described elsewhere (Yabe et al., 2020).

### 2.3 Statistical methods

We used spatial autocorrelation methods involving the global Moran’s I and local Getis-Ord Gi*statistic to assess the spatial patterns in the children’s BLL. These methods are briefly discussed below.

#### a. Test for spatial dependency

The global Moran’s I was implemented in ArcMap 10.5.1(Release, 2012) to test for a general spatial dependency among the BLL in children in Kabwe, i.e. to examine whether high or low levels of BLL show spatial clusters or whether they are scattered in a random pattern.

The global Moran’s I is given by the formula:

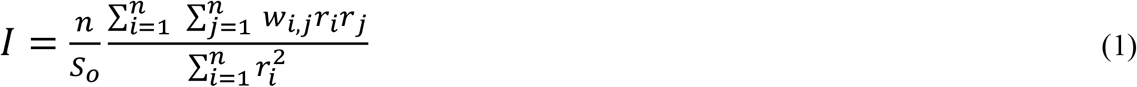

Where *r*_*i*_ is the deviation of the child’s BLL value at area *i* from its mean (*x*_*i*_− *μ*), *w*_*i,j*_ *i*s the spatial weight between area, *i* and *j, n* are the numbers of observations and *S*_*o*_ is the sum of all the spatial weights:

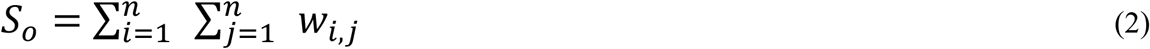

#### b. Hotspot analysis

To identify the spatial location (coordinates) of cluster of high and of low BLL levels (hotspots and coldspots), we used the local Getis-Ord Gi*statistic (Getis and Ord, 2010) We examine the BLL observation with respect to neighbouring BLL observations. An observation with a high value or low value does not necessarily imply a hotspot or a coldspot, respectively, unless it is surrounded by observations with high values (hotspot) or low values (coldspot). Thus, a a large positive Gi*statistic is obtained when the local sum of an observation and its neighbours is larger than the expected local sum indicating clustering of high values. While a small values of the Gi*statistic indicates clustering of low BLL values. In addition we evaluate, whether the Gi*statistic significantly differs from 0, i.e. whether a cluster has a significantly elevated or significantly low BLL level, leading to the definition of hot spots and cold spots, respectively.

. The Gi*statistic is given as:

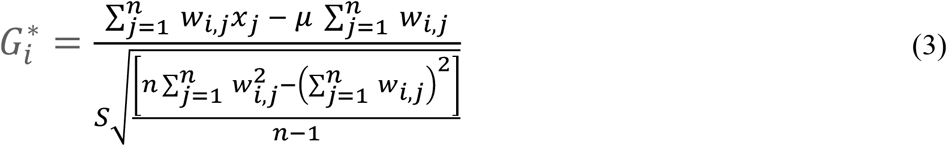

Where *x*_*j*_ is the BLL value for the child in area *j, w*_*i,j*_ is the spatial weight between area *i* and *j, n* is the number of observations, mu is the mean BLL level and S is the standard deviation of x, i.e.:

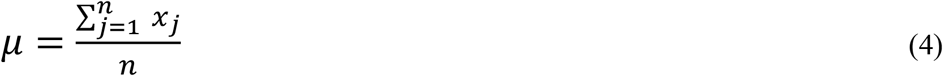

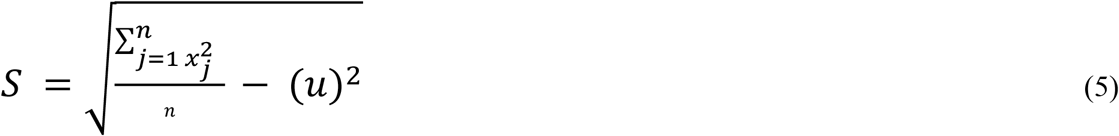

All spatial analyses were performed using ArcMap 10.5.1. All test decisions were based on a significance level of 0.05.

### 2.4 Ethical clearance

The University of Zambia Research Ethics Committee (UNZAREC; REF. No. 012-04-16) approved the study. The Ministry of Health through the Zambia National Health Research Ethics Board and the Kabwe District Medical Office granted further approvals (Yabe et al., 2020). In accordance to ethical guidelines and data protection, until the point of data integration and analysis the location coordinates were stored separately from attribute data, all attribute data was de-identified.

## 3. Results

Table 1 shows the age groups and the distribution of BLL by age groups for our sample. The largest group (47.7%) was the age 0-3 years while the smallest (5%) was the group of 10-15 years old children. The global mean BLL was 30.14 µg/dL and the median 23.75 µg/dL, which is comparable to the numbers reported for the young child by Yabe (Yabe et al., 2020). The distribution of individual BLL ranged from a minimum of 3 µg/dL to maximum of 162 µg/dL. Lowest mean and median BLL (28.7 µg/dL, 21.9 µg/dL) was observed in age group 4-9 years.

**Table 1:**
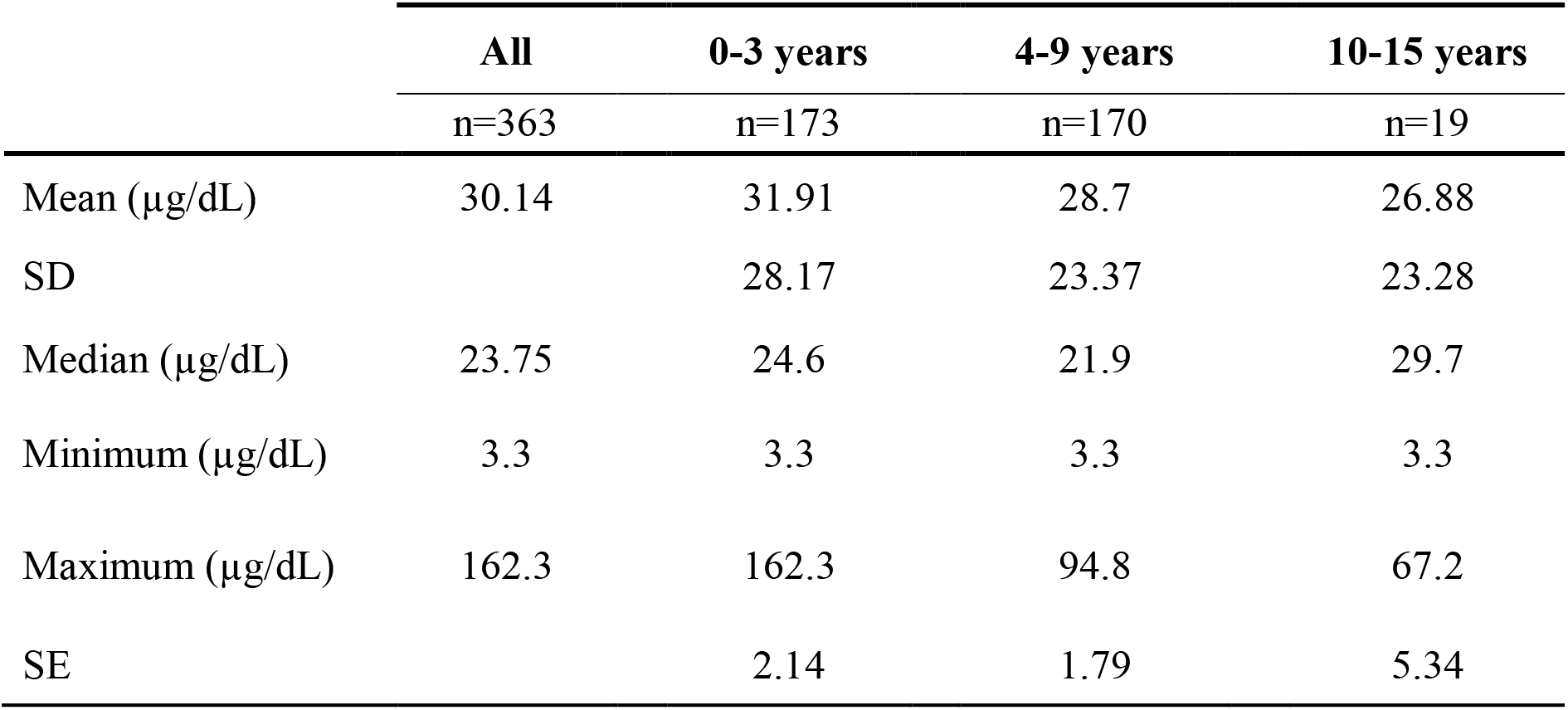
Blood lead level (BLL) distribution

|  | All | 0-3 years | 4-9 years | 10-15 years |
| --- | --- | --- | --- | --- |
|  | n=363 | n=173 | n=170 | n=19 |
| Mean ($\mu\text{g/dL}$) | 30.14 | 31.91 | 28.7 | 26.88 |
| SD |  | 28.17 | 23.37 | 23.28 |
| Median ($\mu\text{g/dL}$) | 23.75 | 24.6 | 21.9 | 29.7 |
| Minimum ($\mu\text{g/dL}$) | 3.3 | 3.3 | 3.3 | 3.3 |
| Maximum ($\mu\text{g/dL}$) | 162.3 | 162.3 | 94.8 | 67.2 |
| SE |  | 2.14 | 1.79 | 5.34 |

**Table 2:** Spatial autocorrelation analysis of blood lead levels (BLL) among the children

| Moran's Index | Expected Index | Variance | Z-Score | p-value |
| --- | --- | --- | --- | --- |
| 0.63 | -0.003 | 0.0006 | 26.1 | <0.001 |

We observe a positive Global Moran’s I (Index) of 0.62 (p-value < 0001) indicating significant spatial clustering of children’s BLL levels in Kabwe. This finding is supported in Figure 2, mapping the BLL values. Red dots indicate areas with high Pb concentration surrounded by other samples with a high concentration (hotspots). Whereas the blue dots indicate areas with low Pb concentration surrounded by other low Pb levels (cold spots) (Wang et al., 2009, Zhang et al., 2008).

**Figure 2.**
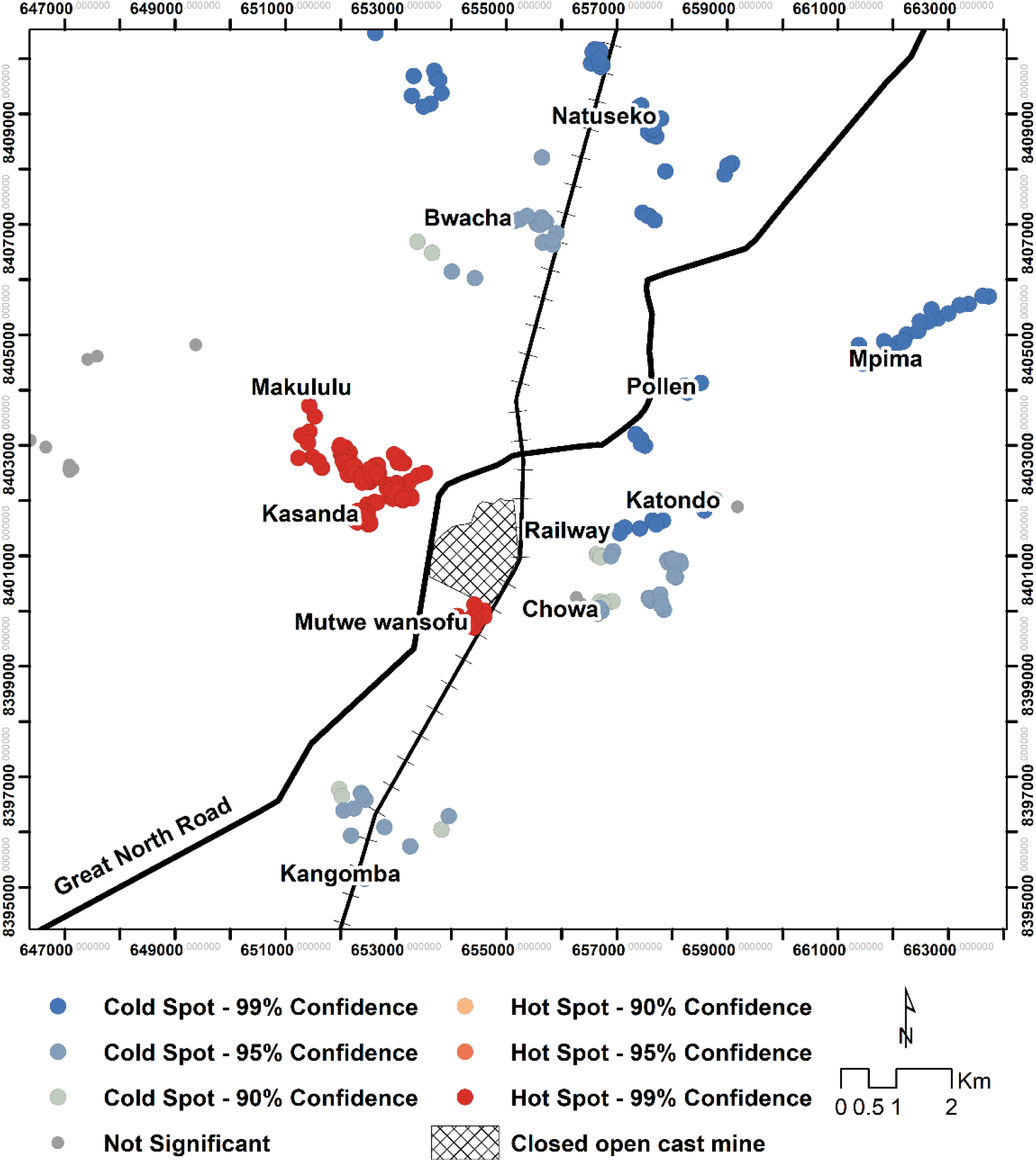
Spatial distribution of blood lead levels among the children in Kabwe, Zambia

We see a clear spatial pattern in the distribution of BLL. Hot spot residential areas are seen on the western side of the Pb open cast mine. These areas include Kasanda, which was a mine residential area for the lowest skilled mine workers and Makululu an informal settlement adjacent to Kasanda. On the south side close to the mine site, Mutwe wansofu is another hot spot area. On the other hand, the northern side is mainly characterised by cold spots. We see Natuseko on the north, Mpima (northeast) and Katondo (east) all being cold spots.

## 4. Discussion

Using secondary data with geo-coordinates, we analysed the spatial autocorrelation of blood lead level (BLL) in Kabwe. Global spatial pattern is particularly important in environmental exposures. The geospatial techniques in this regard help to identify the high/low-exposure areas and this information can serve as a basis for setting up targeted health intervention programs and other environmental remedial measures in affected areas (Oyana and Margai, 2010, Requia et al., 2017).

We found significant spatial patterns in the BLL of children from Kabwe. The clustering of high BLL in specific communities is consistent with the suggestion that source of the metal pollution is historical mining activity at the now abandoned Kabwe open cast Pb-Zn mine. (Yabe et al., 2015a, Ettler et al., 2020).

Wind direction in Kabwe is predominantly east to west. It was observed that the hot spots areas were close to the mine and on the windward side of the mine. Wind blowing top loose soil from the mine site could account for this observed pattern. A study by Yabe et al. (2020) also noted that distance from the mine site had a stronger and bigger negative relationship with BLL in a principal component analysis. Nakayama et al. (2011) in their study also concluded that the source of the pollution in Kabwe was the abandoned Pb-Zn mine.

From the hot spot analysis, we see a clear pattern where hot spot areas lie to the western side, and in close proximity to the mine. These areas include Kasanda and Mutwe Wansomfu. Conversely, we find distinct cold spots areas further away from the mine site (Mpima, Natuseko, Kangombe) and generally more on the eastern side. These identified hotspots are consistent with earlier reported results where children in Kasanda had the highest mean BLL among other communities in Kabwe (Yabe et al., 2015b, Bose-O’Reilly et al., 2018). Contrariwise, our observed cold spots are similar areas earlier reported to have low BLLs in children. Our findings show that the direction from the mine is a major factor explaining the clustering and hot spots of BLL in children in Kabwe. For example, residential areas such as Katondo and Chowa are closer to the mine than Makululu. However, they have not been identified as hotspots as they lie on the eastern side of the mine This backs the hypothesis of the association between the windward side and high BLL. The children in the identified hotspot areas could be getting the lead from the soil as these residential areas are amongst the areas that were reported to have the highest soil lead levels (Bose-O’Reilly et al., 2018, Ettler et al., 2020). The high levels of Pb in soil are due to the fact that the neither the open pit mine, nor the tailing hill were ever properly rehabilitated, and contaminated the areas with Pb since decades. As well, most residential areas lack greenness especially during rainy season very dusty. Children live and play in those dusty surroundings, exposing themselves to lead from dust and soil..

It is also noted that socio-economic factors could also be at play. Historically, Kasanda, Chowa and Luangwa are townships that belonged to the mine. Kasanda on the western side was for the less skilled workers of the mine, while Chowa and Luangwa were for the more skilled workers and expatriates. Makululu is an informal settlement that sprung up from immigrants, mostly less skilled who were searching for job in the mines. Thus, this also backs the hypothesis of windward direction as residential areas for the mining expatriates are unlikely to have been located on the windward side. The exposure of the Kasanda and Makululu communities to lead is likely to be higher due to proximity to the mine, windward location of these residential areas and the socio-economic factors as these are mainly low-income areas.

## 5. Conclusion

The geospatial approach used in the present study has provided vivid insight in spatial patterns of blood lead levels in the children of Kabwe. The study has established clustering effect of BLL and identified hot spot areas. Clearly, the BLL levels are depending on distance and windward direction from the old mine site. Remedial and treatment interventions should consider this, and prioritize these affected communities. The relationship of the observed soil Pb levels and distributions of BLL is yet to be established. Further research is needed to develop a model using lead soil data to predict dangerous childhood Pb exposure. Soil Pb levels are relatively easy and cheap to collect. With a similar spatial analysis on soil levels hot spot areas could be detected, and the progress of interventions could be very well followed and documented. To establish for Kabwe the still unknown attributable fraction of each exposure pathway (inhalation, ingestion) and each source of exposure (soil, air, water, food), spatial modelling would be an ideal tool.

## Data Availability

Data used in this current study is held under confidentiallity as outlined in the ethical approaval. It has since been de-indentified and can be availed when requested.

## 6. Acknowledgements

This work was supported by the CIH^LMU^ Center for International Health and OH-TARGET (One Health Training And Research Global Network) funded by the German Federal Ministry for Economic Cooperation and Development (BMZ) and coordinated by the German Academic Exchange Service (DAAD).

Also by Grants-in-Aid for Scientific Research from the Ministry of Education, Culture, Sports, Science, and Technology of Japan awarded to M. Ishizuka (No. 16H0177906, 18K1984708), S.M.M. Nakayama (No. 16K16197, 17KK0009), and H. Nakata (No. 19K2047209) and the foundation of JSPS Bilateral Open Partnership Joint Research Projects (JPJSBP120209902). We also acknowledge financial support from The Nihon Seimei Foundation, The Japan Prize Foundation, and The Akiyama Life Science Foundation. This research was also supported by JST/JICA, SATREPS (Science and Technology Research Partnership for Sustainable Development; No. JPMJSA1501) and JST aXis (Accelerating Social Implementation for SDGs Achievement).

